# Germline and Tumor Whole Genome Sequencing as a Diagnostic Tool to Resolve Suspected Lynch Syndrome

**DOI:** 10.1101/2020.03.12.20034991

**Authors:** Bernard J. Pope, Mark Clendenning, Christophe Rosty, Khalid Mahmood, Peter Georgeson, Jihoon E. Joo, Romy Walker, Ryan A. Hutchinson, Harindra Jayasekara, Sharelle Joseland, Julia Como, Susan Preston, Amanda B. Spurdle, Finlay A. Macrae, Aung K. Win, John L. Hopper, Mark A. Jenkins, Ingrid M. Winship, Daniel D. Buchanan

## Abstract

**Background:** People who develop mismatch repair (MMR) deficient cancer in the absence of a germline MMR gene pathogenic variant or hypermethylation of the *MLH1* gene promoter in their tumor are classified as having suspected Lynch syndrome (SLS). We applied germline whole genome sequencing (WGS) and targeted and genome-wide tumor sequencing approaches to identify the underlying cause of tumor MMR-deficiency in SLS.

**Methods:** Germline WGS was performed on 14 cancer-affected people with SLS, including two sets of first-degree relatives. Tumor tissue was sequenced for somatic MMR gene mutations by targeted, whole exome sequencing or WGS. Germline pathogenic variants, including complex structural rearrangements and non-coding variants, were assessed for the MMR genes. Tumor mutation burden and mutational signatures.

**Results:** Germline WGS identified pathogenic MMR variants in 3 of the 14 (21.4%) SLS cases including a 9.5Mb inversion disrupting exons 1-7 of *MSH2* in a mother and daughter. Excluding these 3 MMR carriers, tumor sequencing identified at least two somatic MMR gene mutations in 8/11 (72.7%) tumors tested, supporting a non-inherited cause of tumor MMR-deficiency. In the second mother-daughter pair, the combined analysis of germline and tumor by WGS supported a somatic rather than inherited cause of their tumor MMR-deficiency, through presence of double somatic *MSH2* mutations in their respective tumors.

**Conclusion:** Germline WGS of people with SLS improved the identification of Lynch syndrome. When coupled with tumor sequencing, >70% of the people with SLS were resolved as having double somatic MMR mutations and a non-inherited cause for their tumor MMR-deficiency.

## INTRODUCTION

Lynch syndrome (LS) is an autosomal dominantly-inherited disorder caused by germline variants affecting the DNA mismatch repair (MMR) genes *MLH1, MSH2, MSH6*, and *PMS2*^1^. Carriers of pathogenic variants in MMR genes have an increased risk of multiple cancer types, predominantly colorectal cancer (CRC) and endometrial cancer (EC)^2^. A key characteristic of LS-related tumors is DNA MMR-deficiency, evidenced by microsatellite instability (MSI)^3^, and loss of MMR protein expression. However, MMR-deficiency is not unique to LS and can occur when both alleles of an MMR gene are deactivated by somatic changes (such as *MLH1* promoter methylation, loss of heterozygosity (LOH) or mutation). Thus, it is important to distinguish MMR-deficient cancers that arise from LS and those that arise sporadically because people with LS and their relatives can benefit from intensive clinical management and surveillance.

Individuals with a MMR-deficient tumor with no evidence of sporadic aetiology and for whom no germline MMR pathogenic variants can be identified are defined as having “suspected Lynch syndrome” (SLS). This group may represent over 50% of the MMR-deficient CRCs and ECs in population-based studies^4^ and poses significant clinical challenges due to screening of family members that is not based on carrier status. Potential explanations for SLS include: 1) artefactual loss of MMR protein expression or incorrect interpretation of immunohistochemical (IHC) staining^5, 6^, 2) germline pathogenic MMR variants are present but are not identified or are outside of the current testing scope^7, 8^, 3) germline pathogenic variants in non-MMR genes that include the *MUTYH*^9^ and *POLE/POLD1*^10^ genes and 4) unidentified pathologic processes leading to MMR-deficiency^7^. Studies have shown that somatic inactivation of both MMR-gene alleles (“double or biallelic somatic”) is observed in up to 70% of SLS-related cancers^11-14^. Therefore, diagnostic approaches that can resolve the underlying aetiology of SLS are needed for optimal risk assessment and management.

We assessed the utility of germline whole genome sequencing (WGS) and targeted tumor sequencing to determine germline and somatic causes of MMR-deficiency in 14 SLS cases from the Australasian Colorectal Cancer Family Registry (ACCFR)^15, 16^ and the Australian National Endometrial Cancer Study (ANECS)^17^. Additionally, WGS of the tumors in two mother-daughter pairs with SLS was used to estimate tumor-related features such as MSI, tumor mutational burden and somatic mutational signatures, contributing to the diagnosis of an inherited or sporadic aetiology in their families.

## METHODS

### Study participants

Participants were identified from either the population-based or clinic-based recruitment arms of the ACCFR^15, 16^ and from the population-based ANECS^17^ studies and were included in this study if they met the strict inclusion criteria for SLS:

1. one or more MMR-deficient tumors determined by loss of expression of one or more MMR proteins by IHC, *and*
2. where the tumor showed loss of expression of the MLH1 and PMS2 proteins and showed no evidence of *MLH1* promoter methylation in the tumor, *and*
3. had no pathogenic variant detected by germline Sanger sequencing or Multiplex Ligation-dependent Probe Amplification (MLPA) of the MMR gene/s indicated as defective by the pattern of protein loss by IHC.

A family history of cancer meeting the Amsterdam I and II^18^ clinical criteria or Revised Bethesda guidelines^19^ were derived as previously described. Two LS individuals with previously identified pathogenic MMR gene variants were WGS as positive controls: Control sample 1 (C1) carried a pathogenic intronic *MSH2* variant (c.212-478T>G) within a highly repetitive region that has been shown to disrupt splicing^8^; Control sample 2 (C2) carried a 1928 bp deletion of *MSH2* exon 6 (chr2:47640695-47642623) (**Supplementary Figure 1A**). Written informed consent was obtained from all participants to collect blood and tumor samples for research purposes.

**Figure 1.**
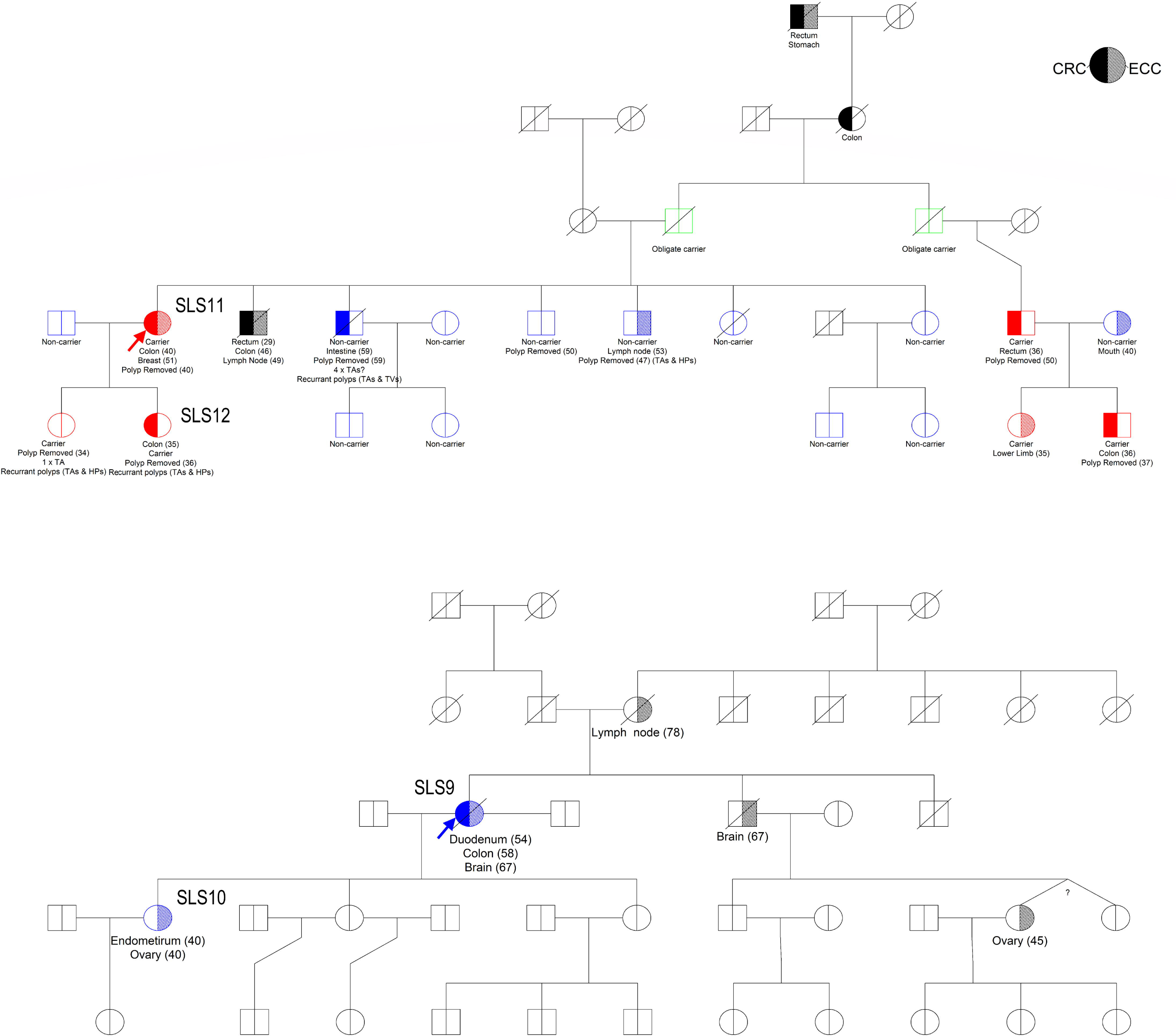
Family pedigrees for two sets of mother-daughter pairs: A) SLS11 and SLS12; and B) SLS9 and SLS10. Probands SLS11 (Family A) and SLS9 (Family B) are indicated with arrows. For Family A an inversion-specific PCR test was used to genotype 14 additional relatives identifying 4 additional carriers (2x CRC-affected, and 1x developed adenomas at 34 years) and two additional obligate carriers (both unaffected).

### MMR molecular testing

All tumors from the study participants were MMR-deficient as determined by IHC staining as previously described^20, 21^. A subset of tumors were tested for MSI using a ten-marker panel as previously described^21^. Tumor methylation of the *MLH1* gene promoter was performed using MethyLight assays as previously described^17, 20, 22^. Testing for the *BRAF* p.V600E somatic mutation was performed on the CRCs and adenomas using a fluorescent allele-specific PCR assay^23^. Germline testing for MMR gene variants was performed using Sanger sequencing and MLPA, including testing for 3′ deletions in *EpCAM* as previously described^17, 20^.

### Germline and Tumor Sequencing

Germline WGS was performed on peripheral blood-derived DNA from all SLS cases and controls (see Supplementary Methods). All SLS-related tumors were screened for somatic mutations in the *MLH1, MSH2, MSH6, PMS2, MLH3, MSH3, PMS1, EXO1*, and *EpCAM* genes using a custom-designed AmpliSeq targeted resequencing assay. A subset of these tumors also had whole genome sequencing through a service provider (see Supplementary Methods).

### Bioinformatic Analysis of Sequencing data

Germline and somatic variant calling and annotation of single nucleotide variants (SNVs), short insertions and deletions (INDELs), Structural Variants (SVs) and Copy Number Variants (CNVs) were detected from the sequencing data using standard techniques (described in Supplementary Methods). All SNVs were assessed using HumanSplicingFinder (HSF) and MaxEntScan for their predicted effect on splicing within the gene/s indicated to be deficient by IHC. Somatic mutational signatures from the set of 30 standard profiles published on the COSMIC website were calculated for each tumor sample using their identified somatic SNVs. Tumor mutation burden (TMB) was calculated as the rate of somatic SNVs per megabase in the coding region of the genome, with TMB >10 considered hypermutated and with TMB >100 considered ultra-hypermutated^24^. Tumor LOH was assessed by comparing the allele fraction of germline variants within tumor samples. Further details regarding the bioinformatic analysis are provided in the Supplementary Methods.

### Candidate Gene Prioritisation

Germline and somatic variants were annotated and prioritised by their intersection with three tiers of genes (**Supplementary Table 1**), with 2kb flanking regions added around the transcription start and stop sites. The highest priority genes (Tier 1, n=5) contained the four MMR genes plus *EpCAM*. Variants intersecting promoter and enhancer regions of the MMR gene indicated to be defective in an individual’s tumor were annotated using promoter and enhancer regions taken from the GeneHancer database^25^. Tier 2 comprised genes assessed by the ClinGen Hereditary Colorectal Cancer and Polyposis Susceptibility Gene Curation Panel to have definitive, strong or moderate evidence supporting an association with hereditary CRC and/or polyposis (*APC, ATM, AXIN2, BLM, BMPR1A, GREM1, MLH3, MUTYH, POLD1, POLE, PTEN, SMAD4, STK11*) or other syndromes with rare manifestation of CRC and/or polyposis (*FLCN, TP53, CDH1*)^26^ or recently reported recessive polyposis genes (*MSH3*^*27*^, *NTHL1*^*28, 29*^) (n=18). Tier 3 contains a curated list of DNA repair genes reported in the literature^30, 31^ (n=265).

**Table 1.**
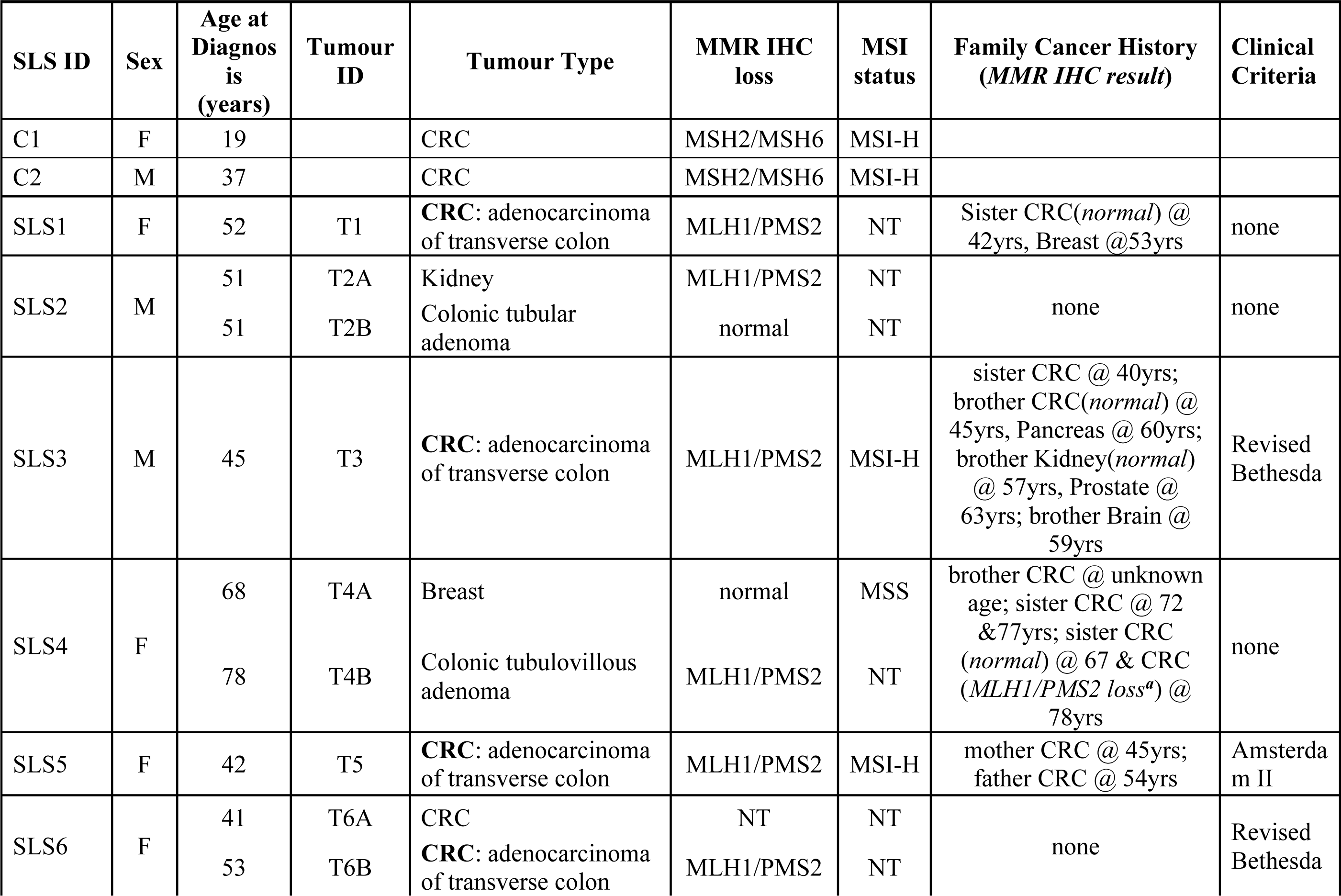
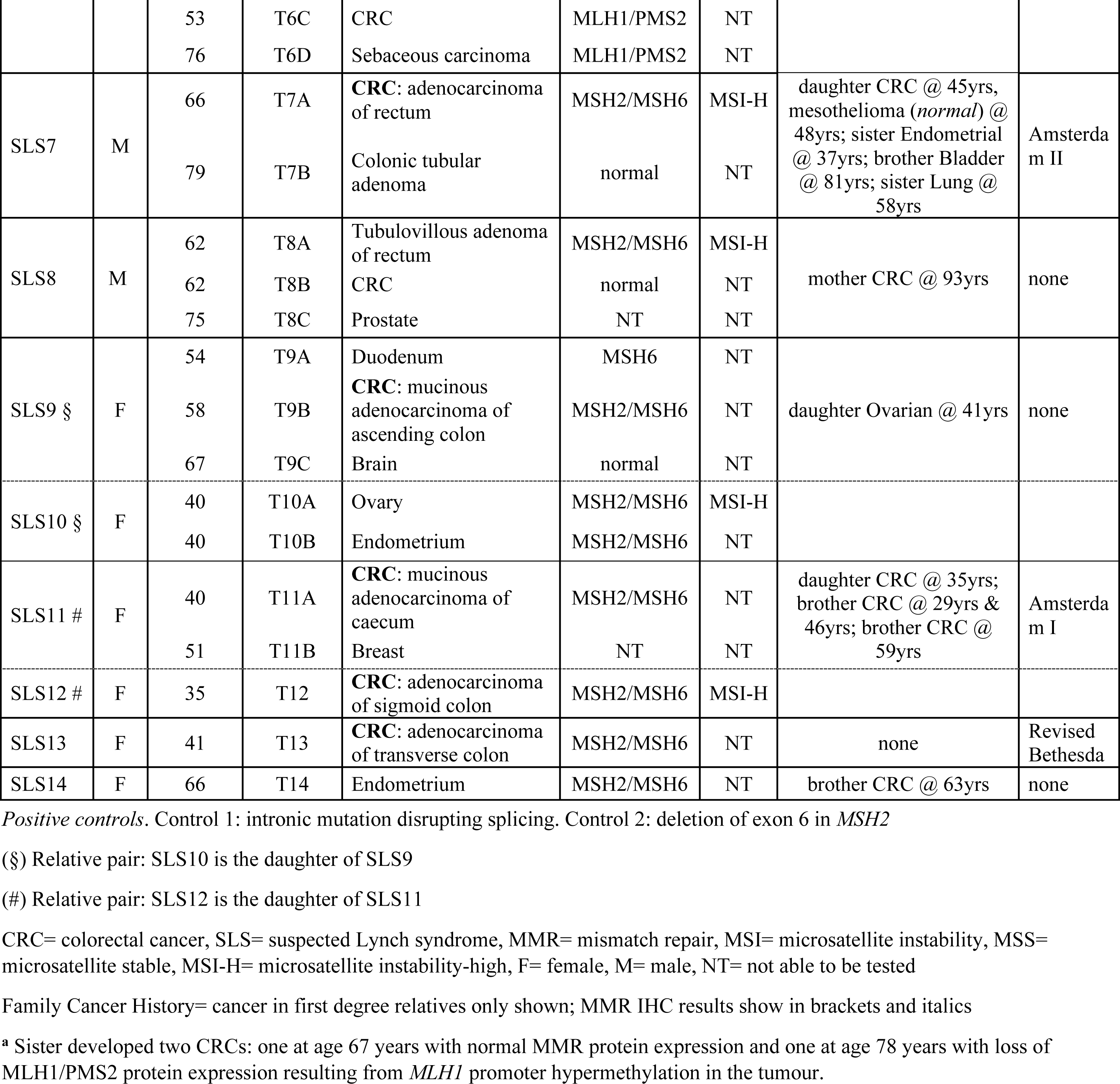
Characteristics of the participants and their tumours that were included in this study.

### Variant Filtering and Prioritisation

Germline SNVs and INDELs identified in Tier 1 or 2 genes were prioritised if they were predicted to have an impact on function (truncating, frameshift and splice site) or nonsynonymous variants resulting in a missense substitution with a CADD score >= 15 or a REVEL score >= 0.6. Tier 3 Germline SNVs and INDELs were prioritised if they were loss of function or high impact variants only. Filtering for population frequency encompassed prioritising variants with a gnomAD allele frequency (AF) <= 2×10^−4^ (except for known pathogenic variants in *MUTYH* NM_001128425.1: c.536A>G p.Tyr179Cys (AF=1.5×10^−3^) and NM_001128425.1: c.1187G>A. p.Gly396Asp (AF=3×10^−3^) and in *NTHL1* NM_002528.5: c.268C>T p.Gln90Ter (AF=1.4×10^−3^). See Supplementary Methods for details on threshold selection criteria. All germline SNVs and INDELs were annotated with ClinVar clinical classifications, and those with at least 2-star review status (criteria provided, multiple submitters, no conflicts) classified as benign or likely-benign were excluded. Prioritised germline variants were validated by Sanger sequencing and tested in relatives with a DNA sample available (Supplementary Methods).

High confidence somatic SNVs and INDELs called by at least 2/3 callers with a variant allele frequency (VAF) >= 0.10 were retained for further analysis. For targeted Ampliseq tumor sequencing, SNVs and INDELs were called with a VAF of >= 0.07. For Tier 1 genes, coding SNVs and INDELS with an InSiGHT classification >= 3 were prioritised. Intronic variants predicted to disrupt splicing or intersecting a promoter or enhancer region of the MMR gene indicated as defective by IHC were also prioritised. For Tier 2 and 3 genes, only loss of function variants were considered unless genes were known to harbour somatic hotspots (e.g. the exonuclease domain in *POLE*).

## RESULTS

The characteristics of the 14 study participants with SLS are described in **Table 1**. Age at diagnosis of MMR-deficient tumor ranged from 35 to 78 years (median=52 years) and 10/14 (71%) were female. A family history of cancer meeting Amsterdam I or II clinical criteria was observed for 3/14 (21.4%) SLS cases. Two sets of mother-daughter pairs (SLS9 and SLS10, and SLS11 and SLS12) were included in the cohort (**Table 1, Figure 1A and 1B**). Seven tumors showed loss of MLH1/PMS2 expression and were negative for the *BRAF*^V600E^ somatic mutation in addition to being negative for tumor *MLH1* gene promoter hypermethylation.

### Germline Whole Genome Sequencing Analysis

Germline variants retained after filtering and prioritisation are provided in **Table 2**. Of all 14 SLS individuals screened by WGS, three carried predicted pathogenic germline MMR variants including a *MLH1* missense variant c.1958T>G, p.Leu653Arg in SLS6, classified as a Class 4 (likely pathogenic) by InSiGHT, and a 9.5 Mb inversion encompassing exons 1-7 of *MSH2* (chr2:38121107-chr2:47669532) in mother-daughter pair of SLS11 and SLS12 (**Supplementary Figure 1B 1C**). An inversion-specific PCR test (Supplementary Methods) was used to genotype 14 additional relatives identifying 4 additional carriers (2x CRC-affected, and 1x developed adenomas at 34 years) and two additional obligate carriers (both unaffected) (**Figure 1A**). Sanger sequencing of the inversion-specific PCR product confirmed that this is the same inversion as previously reported^32^.

**Table 2.**
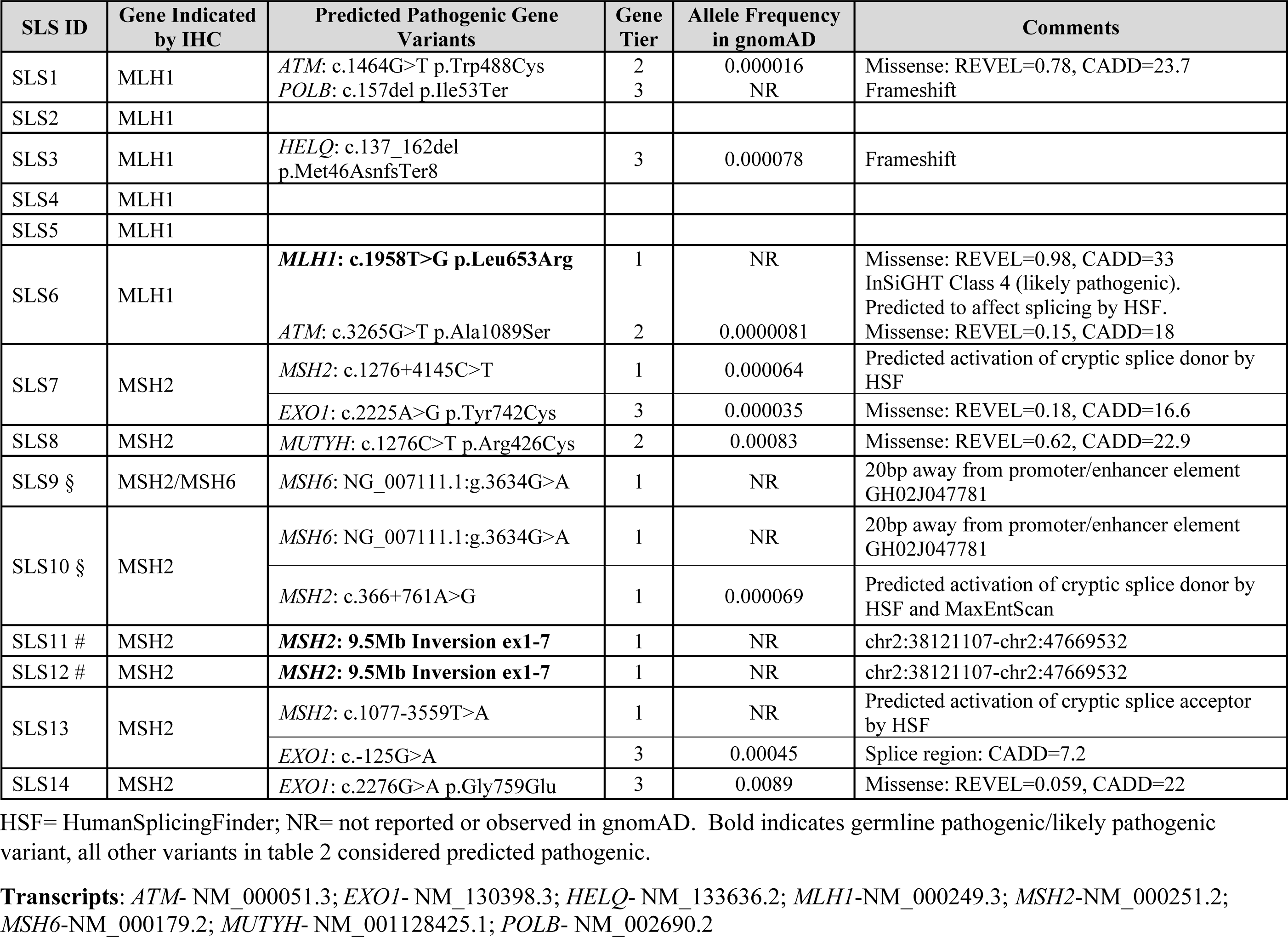
Predicted germline pathogenic variants identified from whole genome sequencing of the 14 suspected Lynch syndrome cases.

| SLS ID | Gene Indicated by IHC | Predicted Pathogenic Gene Variants | Gene Tier | Allele Frequency in gnomAD | Comments |
| --- | --- | --- | --- | --- | --- |
| SLS1 | MLH1 | *ATM*: c.1464G>T p.Trp488Cys<br>*POLB*: c.157del p.Ile53Ter | 2<br>3 | 0.000016<br>NR | Missense: REVEL=0.78, CADD=23.7<br>Frameshift |
| SLS2 | MLH1 |  |  |  |  |
| SLS3 | MLH1 | *HELQ*: c.137_162del p.Met46AsnfsTer8 | 3 | 0.000078 | Frameshift |
| SLS4 | MLH1 |  |  |  |  |
| SLS5 | MLH1 |  |  |  |  |
| SLS6 | MLH1 | ***MLH1*: c.1958T>G p.Leu653Arg** | 1 | NR | Missense: REVEL=0.98, CADD=33<br>InSiGHT Class 4 (likely pathogenic).<br>Predicted to affect splicing by HSF. |
|  |  | *ATM*: c.3265G>T p.Ala1089Ser | 2 | 0.0000081 | Missense: REVEL=0.15, CADD=18 |
| SLS7 | MSH2 | *MSH2*: c.1276+4145C>T | 1 | 0.000064 | Predicted activation of cryptic splice donor by HSF |
|  |  | *EXO1*: c.2225A>G p.Tyr742Cys | 3 | 0.000035 | Missense: REVEL=0.18, CADD=16.6 |
| SLS8 | MSH2 | *MUTYH*: c.1276C>T p.Arg426Cys | 2 | 0.00083 | Missense: REVEL=0.62, CADD=22.9 |
| SLS9 § | MSH2/MSH6 | *MSH6*: NG_007111.1:g.3634G>A | 1 | NR | 20bp away from promoter/enhancer element<br>GH02J047781 |
| SLS10 § | MSH2 | *MSH6*: NG_007111.1:g.3634G>A | 1 | NR | 20bp away from promoter/enhancer element<br>GH02J047781 |
|  |  | *MSH2*: c.366+761A>G | 1 | 0.000069 | Predicted activation of cryptic splice donor by HSF and MaxEntScan |
| SLS11 # | MSH2 | ***MSH2*: 9.5Mb Inversion ex1-7** | 1 | NR | chr2:38121107-chr2:47669532 |
| SLS12 # | MSH2 | ***MSH2*: 9.5Mb Inversion ex1-7** | 1 | NR | chr2:38121107-chr2:47669532 |
| SLS13 | MSH2 | *MSH2*: c.1077-3559T>A | 1 | NR | Predicted activation of cryptic splice acceptor by HSF |
|  |  | *EXO1*: c.-125G>A | 3 | 0.00045 | Splice region: CADD=7.2 |
| SLS14 | MSH2 | *EXO1*: c.2276G>A p.Gly759Glu | 3 | 0.0089 | Missense: REVEL=0.059, CADD=22 |

HSF= HumanSplicingFinder; NR= not reported or observed in gnomAD. Bold indicates germline pathogenic/likely pathogenic variant, all other variants in table 2 considered predicted pathogenic.
**Transcripts:** *ATM*- NM\_000051.3; *EXO1*- NM\_130398.3; *HELQ*- NM\_133636.2; *MLH1*-NM\_000249.3; *MSH2*-NM\_000251.2; *MSH6*-NM\_000179.2; *MUTYH*- NM\_001128425.1; *POLB*- NM\_002690.2

### Somatic MMR gene analysis

Targeted tumor sequencing of the MMR genes was completed for 14 tumors from 13 SLS cases (T4A failed testing, **Table 1**) where 11 of the tested tumors occurred in the colon or rectum. A subset of five tumors were selected for additional WGS (n=4) or WES (n=1) from individuals where no pathogenic germline variants were identified. In the genes indicated to be defective by IHC we identified at least two somatic mutations in 9/14 (64.3%) tumors, a single somatic mutation in 3/14 (21.4%) tumors and no somatic mutations in 2/14 (14.3%) tumors (**Table 3**). No somatic SVs or CNVs intersecting with the four MMR genes were detected.

**Table 3.** Somatic MMR gene mutations identified from targeted MMR gene testing and from whole genome or exome sequencing of the suspected Lynch syndrome tumours.

| SLS ID | Age at Dx (yrs) | Tumour ID | Tumour Type | MMR IHC loss | MMR tumour classification | MMR somatic mutations (VAF) |
| --- | --- | --- | --- | --- | --- | --- |
| SLS1 | 52 | T1 | CRC | MLH1/PMS2 | Double somatic | MLH1: c.879C>G p.Tyr293Ter (0.11)<br>MLH1: c.1975C>T p.Arg659Ter (0.10) |
| SLS3 | 45 | T3 | CRC | MLH1/PMS2 | Single somatic | MLH1: c.332C>T p.Ala111Val (0.34) |
| SLS4 | 78 | T4B | Tubulovillous adenoma | MLH1/PMS2 | Double somatic | MLH1: c.1036C>T p.Gln346Ter (0.07)<br>MLH1: c.812C>T p.Ser271Phe (0.15)<br>MLH1: c.865C>T p.His289Tyr (0.12) |
| SLS5 | 42 | T5 | CRC<sup>‡</sup> | MLH1/PMS2 | Double somatic | MLH1: c.1989+1G>A (0.63)<br>MLH1: LOH |
| **SLS6<sup>a</sup>** | 53 | T6B | CRC | MLH1/PMS2 | Double somatic | MLH1: c.1348dup p.Asp450GlyfsTer29 (0.16)<br>MLH1: c.1834G>A p.Val612Ile (0.07) <sup>c</sup> |
| SLS7 | 66 | T7A | CRC | MSH2/MSH6 | Double somatic | MSH2: c.1351C>T p.Gln451Ter (0.11)<br>MSH2: c.1204C>T p.Gln402Ter (0.09) |
| SLS8 | 62 | T8A | Tubulovillous adenoma | MSH2/MSH6 | Double somatic | MSH2: c.859G>T p.Gly287Ter (0.11)<br>MSH2: c.2465_2466delinsAG p.Cys822Ter (0.11) |
| SLS9 § | 54 | T9A | Duodenum* | MSH6 | none | No somatic MMR gene variants detected |
| SLS9 § | 58 | T9B | CRC* | MSH2/MSH6 | Double somatic | MSH2: c.1915C>T p.His639Tyr (0.25)<br>MSH2: c.942+17_942+29del (0.57) <sup>d</sup><br>MSH2: c.1387-2768C>G (0.58) <sup>e</sup><br>MSH2: c.1662-457T>G (0.4) <sup>f</sup><br>MSH6: c.3261del p.Phe1088SerfsTer2 (0.86) |
| SLS10 § | 40 | T10A | Ovary* | MSH2/MSH6 | Double somatic | MSH2: c.308del p.Tyr103LeufsTer71 (0.38)<br>MSH2: c.1277-2187C>G (0.22) <sup>f</sup><br>MSH2: c.1510+1281G>A (0.11) <sup>f</sup><br>MSH2: c.943-912_943-909del (0.7) <sup>e</sup><br>MSH6: c.3261dup p.Phe1088LeufsTer5 (0.5) |
| **SLS11 <sup>#b</sup>** | 40 | T11A | CRC | MSH2/MSH6 | none | No somatic MMR detected |
| **SLS12 <sup>#b</sup>** | 35 | T12 | CRC | MSH2/MSH6 | Single somatic | MSH2: c.2459-12A>G (0.28) |
| SLS13 | 41 | T13 | CRC* | MSH2/MSH6 | Single somatic | MSH2: c.2006-3T>G (0.33) |
| SLS14 | 66 | T14 | Endometrium | MSH2/MSH6 | Double somatic | MSH2: c.1276+2delT (0.37)<br>MSH2: c.2131C>T p.Arg711Ter (0.28) |

Transcripts: *MLH1*-NM\_000249.3; *MSH2*-NM\_000251.2; *MSH6*-NM\_000179.2
Kidney cancer (T2A) showing loss of MLH1/PMS2 expression from SLS2 had insufficient DNA to repeat a failed test result.
Relative pairs: (§) SLS10 is the daughter of SLS9 and (#) SLS12 is the daughter of SLS11
(\*) also tested using whole genome sequencing.
(<sup>¥</sup>) also tested using whole exome sequencing.
<sup>a</sup> Carrier of germline *MLH1* p.Leu653Arg missense variant classified as Class 4 (likely pathogenic).
<sup>b</sup> Carrier of a germline 9.5Mb inversion encompassing exons 1-7 of *MSH2* (chr2:38121107-chr2:47669532).
<sup>c</sup> Variant classified in ClinVar as variant of uncertain significance with 2-star rating.
<sup>d</sup> Intronic deletion called by only 1/3 somatic variant callers used.

We tested two tumors from SLS9 (T9A-duodenal and T9B-CRC) by targeted and WGS. For T9A, IHC indicated loss of MSH6, however no MMR gene variants were found that might disrupt *MSH6* in that tumor (**Table 3**). For the T9B, demonstrating loss of MSH2/MSH6, multiple somatic *MSH2* mutations were observed, including somatic variants predicted to disrupt splicing through activation of both cryptic splicing donor and acceptor sites, plausibly related to the somatic inactivation of both alleles of that gene (**Table 3**). Similarly, the ovarian cancer (T10A) from SLS10 showed loss of MSH2/MSH6 expression where the targeted and WGS tumor sequencing identified multiple somatic *MSH2* variants (**Table 3**).

WES was performed on the CRC tumor T5. The first 50 Mb of the p-arm of chromosome 3, containing *MLH1*, showed clear evidence of LOH (**Figure 2A**). Additionally, we identified a *MLH1*:c.1989+1G>A variant with high VAF in both the targeted (79%) and WES (58%) analysis of the tumor. The combination of this somatic variant and LOH suggests biallelic *MLH1* inactivation in the tumor and is concordant with our IHC results (**Table 3**). None of the other four tumors with WGS showed evidence of LOH (**Figure 2**).

**Figure 2.**
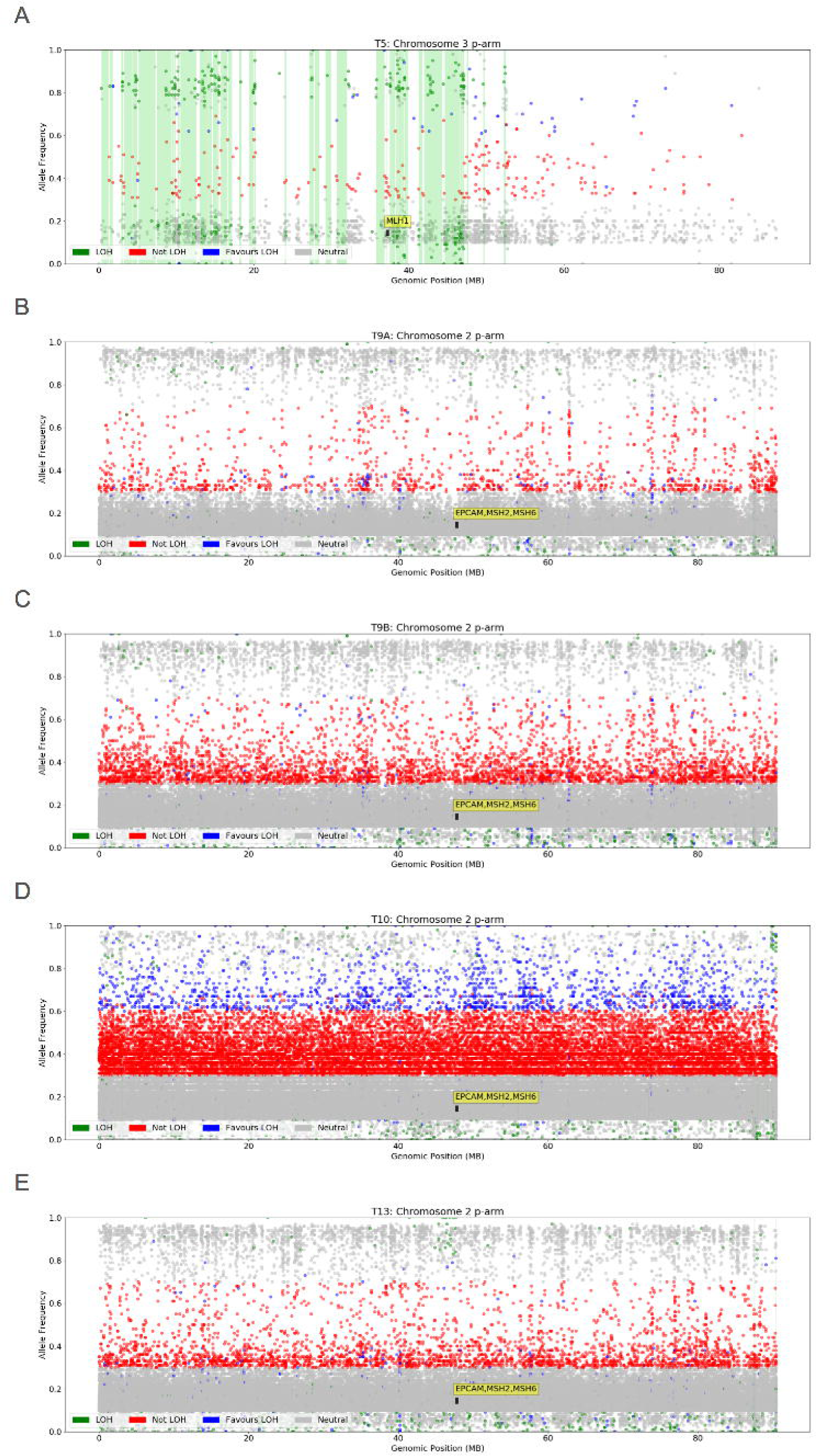
Plots (A-E) depict the presence or the absence of loss of heterozygosity (LOH) in each tumor across the first 90 million bases of: (A) the p-arm of chromosome 3, which encompasses the *MLH1* gene; and (B-E) the p-arm of chromosome 2, which encompasses the *EpCAM, MSH2* and *MSH6* genes. Green vertical bars indicate regions where the variant allele fraction of germline variants within tumor samples deviates from the germline state and thus provides strong evidence for LOH. Tumor sample T5 showed loss of *MLH1* via IHC (**Table 1**) and demonstrated significant regions of LOH surrounding the *MLH1* gene. Tumor sample T9A showed loss of *MSH6* via IHC (**Table 1**) and tumor samples T9B, T10 and T13 showed loss of *MSH2* via IHC (**Table 1**), however none of these tumors demonstrated evidence for LOH in the regions surrounding these genes.

### Tumor Whole Genome and Exome Sequencing Analysis

High impact somatic mutations in genes from Tiers 2 and 3 were identified in each of the five tumors with WGS/WES (**Table 4**). Notably, the three CRCs tested (T5, T9B, T13) had stop-gain mutations in *APC* (**Table 4**). The ovarian tumor (T10A) had a missense mutation in the DNA polymerase gene *POLD1*:p.Glu318Lys, which is situated at the exonuclease active site within the encoded protein. In addition, we investigated features associated with tumor MMR-deficiency namely somatic mutational signatures (signatures 6, 14, 15, 20 and 26), tumor mutational burden (TMB) and MSI, estimated from the sequencing data. The somatic mutational signatures are shown in **Figure 3**, where for four of the five tumors (T5, T9B, T10A, T13) we observed signatures 6, 20 and 26, ranging in contribution from 4% to 69%, supporting tumor MMR-deficiency status determined by MMR IHC (**Table 4**). Only the ovarian tumor (T10A) showed significant contributions from signatures 20 and 14, both of which are associated with MMR-deficiency and defective polymerase proofreading. The duodenal tumor (T9A) did not exhibit any MMR-deficiency associated signatures despite initial MMR IHC testing describing loss of MSH6 protein expression. The MSI status derived from WGS/WES by MSIsensor predicted four of the five tumors tested to be MSI-H, where the duodenal tumor T9A was predicted to be microsatellite stable (MSS) (**Table 4**). Four tumors (T5, T9B, T10A, and T13) exhibited high TMBs consistent with a hypermutator phenotype (or ultra-hypermutator in the case of the ovarian tumor T10A) where the duodenal tumor T9A was not considered to be hypermutated (**Table 4**).

**Table 4.** Tumour characteristics derived from whole genome sequencing and whole exome sequencing analysis of five suspected Lynch syndrome related tumours.

| SLS ID | Tumour ID | MMR-related Mutational Signatures (%) | Other Mutational Signatures (%) | MSI-Sensor | Tumour Mutational Burden (TMB) | Somatic mutations in Tier 2/3 genes (VAF) | Tier |
| --- | --- | --- | --- | --- | --- | --- | --- |
| SLS5 | T5-CRC | 6 (76)<br>26 (4) | 1 (17) | 44.3%<br>(MSI-H) | 38.3 | **APC:** p.Gly132Ter (0.21)<br>**BRCA2:** p.Lys1691AsnfsTer15 (0.19) | 2<br>3 |
| SLS9 § | T9A-Duodenal | none | 30 (51)<br>8 (15)<br>16 (11)<br>2 (10) | 0.06%<br>(MSS) | 8.6 | **MSH3:** c.3001-1G>A (0.24) | 2 |
| SLS9 § | T9B-CRC | 6 (17)<br>26 (9) | 1 (34)<br>8 (10)<br>12 (8)<br>9 (8)<br>25 (7) | 12.0%<br>(MSI-H) | 35.0 | **APC:** p.Arg876Ter (0.3)<br>**MSH3:** p.Lys383ArgfsTer32 (0.45)<br>**BRCA2:** p.Asn1287IlefsTer6 (0.33)<br>**POLD3:** p.Arg300GlyfsTer5 (0.36)<br>**BLM:** p.Asn515MetfsTer16 (0.42)<br>**RPA1:** c.1746+1G>A (0.28) | 2<br>2<br>3<br>3<br>3<br>3 |
| SLS10 § | T10A-Ovarian | 20 (48) | 14 (33)<br>12 (8) | 6.4%<br>(MSI-H) | 117.7 | **POLD1:** p.Glu318Lys (0.47)<br>**POLE:** p.Arg762Trp (0.43)<br>**POLE:** p.Ser80Gly (0.48)<br>**ATM:** p.Gln1117Ter (0.33)<br>**TELO2:** p.Val402ArgfsTer13 (0.5)<br>**LIG1:** p.Cys719Ter (0.44) | 2<br>2<br>2<br>2<br>3<br>3 |
| SLS13 | T13-CRC | 26 (7)<br>6 (7) | 1 (28)<br>30 (19)<br>8 (12)<br>14 (7)<br>12 (6) | 4.1%<br>(MSI-H) | 19.7 | **APC:** p.Arg805Ter (0.43)<br>**BMPR1A:** p.Asn420ThrfsTer16 (0.21)<br>**AXIN2:** p.Gly665AlafsTer24 (0.33)<br>**AXIN2:** p.Arg399Ter (0.41)<br>**EXO1:** p.Arg668Ter (0.26)<br>**PMS1:** p.Glu543Ter (0.2)<br>**HELQ:** p.Lys561ArgfsTer8 (0.38) | 2<br>2<br>2<br>2<br>3<br>3<br>3 |

Somatic mutational signatures were computed using the method described by DeconstructSigs, which estimates the relative contributions of each mutational process in a single tumour sample from the set of 30 standard profiles established by the Wellcome Trust Sanger Institute (WTSI) Mutational Signature Framework (Version 2) as previously described.
Microsatellite instability (MSI) was assessed computationally on the tumour samples using MSIsensor 0.5 using thresholds of $>3.5$ considered to be high levels of MSI (MSI-H), between 1 and 3.5 considered to be low levels of MSI (MSI-L) and $<1$ stable with regards to MSI (MSS).
Tumour Mutational Burden (TMB): derived from the number of somatic SNVs and somatic INDELs per megabase (Mb) in the coding region of the genome where tumours with TMB $>10$ were considered hypermutated and with TMB $>100$ were considered ultra-hypermutated.

**Figure 3.**
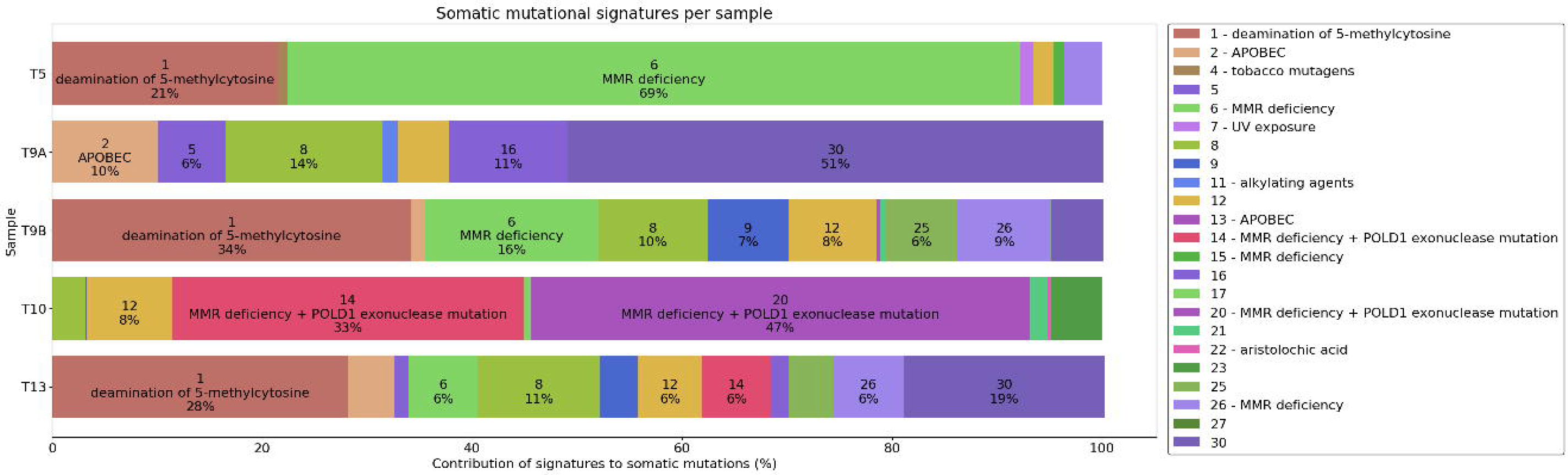
The somatic mutational signature components for tumor samples T5, T9A, T9B, T10 and T13 indicated by the signature number, suggested aetiology and the percentage contribution to the overall mutational composition for each sample.

## DISCUSSION

Germline WGS was applied to 14 people classified as SLS to determine whether an investigation of the DNA MMR genes beyond current conventional testing approaches could improve the detection of germline MMR gene pathogenic variants. Our WGS analysis identified germline pathogenic MMR variants in 3 of the 14 (21.4%) SLS cases, variants which had been previously missed by conventional testing approaches. In addition, we analysed at least one MMR-deficient tumor from 13 SLS cases (1 tumor failed) for somatic mutations within the MMR genes using targeted sequencing and WGS or WES. We identified at least two somatic mutations in the MMR gene indicated as defective by IHC in 9/14 (64.3%) tumors tested, increasing to 72.7% (8/11) when tumors from the three identified carriers of germline pathogenic variants were excluded. Our findings add to the growing number of studies that have shown that double somatic MMR mutations are a common cause of MMR-deficiency in SLS^11-^13, 33^-36^. Furthermore, the application of tumor WGS/WES to the subset of five SLS tumors enabled the determination of additional features associated with defective MMR including somatic mutational signatures 6, 20 and 26, MSI-high status, and high TMB, providing further evidence of a MMR-deficient tumor phenotype and allaying concerns over a false-positive MMR IHC result, although studies validating this approach are needed.

Germline WGS analysis of the mother-daughter pair SLS11 and SLS12, each with a MSH2/MSH6 deficient CRC, identified an inversion encompassing exons 1-7 of *MSH2*. This inversion was first reported in the literature by Wagner *et al* in 2002^37^ and later reported by Rhees *et al*^*32*^ in 2014 and by Mork *et al*^*38*^ in 2017 using various methods, although none employing WGS. Targeted sequencing of the MMR genes in the two corresponding CRCs (T11A and T12) revealed a single predicted pathogenic somatic *MSH2* gene mutation in T12 but not in T11A. More extensive analysis of the T11A CRC using WGS may reveal a “second hit” in *MSH2*.

A second mother-daughter pair (SLS9 and SLS10) underwent germline and tumor WGS (**Figure 1B**). No germline coding variants affecting *MSH2* or *MSH6* were identified. In addition to exonic variants, WGS enabled the investigation of intronic and promoter/enhancer variants in the MMR genes. A germline *MSH6*: NG_007111.1:g.3634G>A variant that resided 20bp upstream from the *MSH6* promoter/enhancer element GH02J047781 (the highest scoring element in the GeneHancer database for *MSH6*) was identified in both individuals. This variant was not observed in gnomAD, highlighting its rarity. Further segregation of this variant in relatives was not possible. Three tumors from these two individuals (T9A, T9B, and T10A) underwent both targeted MMR sequencing and WGS. T9-CRC and T10A-ovarian cancer both acquired somatic frameshift mutations within a coding microsatellite of *MSH6*. Although these somatic *MSH6* mutations may represent the “second hit” subsequent to the abovementioned germline variant, it is also possible that this mutation may be a consequence of MMR-deficiency, rather than a cause. Somatic expansion of this repeat in *MSH6* exon 5 has been previously reported^39^. Two of the three tumors tested for in these people showed loss of MSH2/MSH6 expression suggesting the defect lies in *MSH2*. A rare germline *MSH2*:c.366+761A>G variant predicted by HSF and MaxEntScan to activate a cryptic splice donor site was identified in SLS10 but not in the mother (SLS9). The T9B-CRC and T10A-ovarian cancer both harboured multiple somatic *MSH2* mutations that could potentially inactivate both alleles of *MSH2*, supporting a sporadic aetiology for both these tumors. The T9A-duodenal tumor in the mother showed solitary loss of MSH6 expression although no somatic *MSH2* or *MSH6* mutations were identified.

Four out of the five tumors tested by WGS/WES (T5, T9B, T10A and T13) demonstrated high proportions of mutational signatures 6, 20 and 26, (**Figure 3**), associated with defective DNA MMR^40^. Additionally, the TMB for these four tumors indicated hypermutation and high levels of MSI were predicted by MSIsensor. These findings provide mechanistic evidence of defective MMR in these tumors, consistent with the IHC results. For the duodenal tumor T9A, no MMR-deficiency related somatic mutational signatures were observed, the tumor was predicted to be MSS, and the TMB was <10 mutations/Mb. These results suggest that T9A does not have defective DNA MMR despite the loss of MSH6 indicated by IHC, representing a possible false positive IHC result. Therefore, SLS9 was classified as having one MMR-deficient tumor *with* functional evidence of defective MMR (T9B-CRC) and one MMR-deficient tumor *without* functional evidence of defective MMR (T9A-duodenal), the latter is not suggestive of a LS aetiology.

The ovarian cancer T10A demonstrated strong somatic mutational signatures 20 (47%) and 14 (33%) and was ultra-hypermutated. A recent study has demonstrated that tumors exhibiting both somatic mutational signatures 20 and 14 and an ultra-hypermutation phenotype are associated with defective MMR and (clonal) exonuclease domain mutations in the DNA polymerase gene *POLD1*, where the *POLD1* somatic mutation precedes the loss of MMR^41^. Concordant with the somatic mutational signature profile, a somatic mutation in the exonuclease domain of *POLD1* was observed together with multiple predicted somatic mutations in *MSH2*. These findings suggest a somatic aetiology underlying the MMR-deficiency in T10A. The combined germline and somatic WGS findings for the mother-daughter pair SLS9 and SLS10 argue against a diagnosis of LS for these individuals.

Several studies have shown that double somatic mutations (including LOH), resulting in biallelic MMR-gene inactivation, are a common cause of MMR-deficiency in SLS^11-13, 33-36^. Excluding both mother-daughter relative pairs and the *MLH1* pathogenic variant carrier, six of the eight remaining tumors/SLS cases (SLS1, SLS4, SLS5, SLS7, SLS8 and SLS14) were classified as “double somatics”. This suggests a somatic cause of their tumor MMR-deficiency and is supported by an absence of germline predicted pathogenic MMR variants in five out of these six individuals (SLS1, SLS4, SLS5, SLS8, and SLS14). Additionally, the double somatic cases SLS4, SLS7 and SLS8 developed a second or third primary tumor that showed normal retained expression of the MMR proteins by IHC. This is unlike LS, where synchronous or metachronous tumors develop concordant loss of MMR protein expression. Of interest, five out of the seven double somatic SLS cases had a first-degree relative who developed a LS-related cancer. A recent study by Pearlman *et al*^14^ showed that people with LS who developed CRC were 15 times more likely to have a family cancer history meeting Amsterdam II criteria and 6 times more likely to develop multiple LS-associated cancers compared with people who developed MMR-deficient CRC resulting from double somatic MMR mutations. In our study, two SLS cases with double somatic CRCs (SLS5 and SLS7) had a family history fulfilling the Amsterdam II criteria and three developed multiple tumors (SLS4, SLS7, SLS8). This suggests that family cancer history and multiple synchronous or metachronous tumors, while good predictors of LS, do not exclude a somatic aetiology for tumor MMR-deficiency.

Germline pathogenic variants within the exonuclease domain of *POLE* or *POLD1* genes or biallelic mutations within the *MUTYH* gene may indirectly lead to tumor MMR-deficiency^9, 42,43^. We identified a single SLS carrier (SLS8) heterozygous for *MUTYH*:p.Arg426Cys predicted pathogenic missense variant. No evidence of a second germline *MUTYH* mutation was found, however the corresponding tumor (T8A) had two somatic loss of function mutations in *MSH2*. A previous analysis of the role of germline *MUTYH* mutations in SLS showed a significant association for biallelic *MUTYH* carriers but not for monoallelic carriers^42^, further supporting a somatic aetiology for this MMR-deficient tumor phenotype. Recent studies of SLS have identified germline bioinformatically predicted pathogenic variants in *MUTYH*^33, 35^, *EXO1*^44^, *POLD1*^44^ genes as well as *BUB1*^33^, *SETD2*^33^, *FAN1*^33^, *RFC1*^44^, *RPA1*^44^, *MLH3*^44^, *MSH3*^35^, *AXIN1*^35^, and *AXIN2*^35^. We identified two bioinformatically predicted pathogenic missense variants in the Tier 2 gene *ATM* in two individuals, although one of these occurred in SLS6 who carried the *MLH1* p.Leu653Arg pathogenic variant. In SLS7, SLS13 and SLS14 predicted pathogenic variants were found in *EXO1* (Exonuclease 1), which plays an important role in MMR via strand excision^45^. Of interest, the carrier of *EXO1*:c.-125G>A variant (SLS15), predicted by VEP to be in a potential splice region, also had a somatic mutation in *EXO1*:p.Arg668Ter.

This study has several strengths. The germline WGS approach enabled the detection of complex SVs, identifying the inversion of *MSH2* exons 1-7, and the detection of MMR variants in promoter/enhancer regions and intronic variants predicted to affect splicing. Current approaches may miss these types of variants due to incomplete genome coverage, difficulties with low DNA complexity, and challenges identifying complex mutations with commonly used sequencing technologies. Germline WGS was able to detect a previously identified deep intronic pathogenic variant in control sample 1 (C1) within a highly repetitive intronic region of the *MSH2* gene^8^. This variant was predicted to potentially disrupt splicing by both HSF and MaxEntScan, validating the approach employed in this study. We also detected rare intronic variants that are predicted to disrupt splicing in the likely defective MMR gene in three germline samples and detected a rare germline *MSH6* promoter region variant shared by a mother and daughter. Validation of the effect of these bioinformatically prioritised variants is challenging and is an area where further research is needed. The application of targeted MMR gene sequencing to all but one of the MMR-deficient tumors from the SLS cases enabled the detection of double somatic MMR mutations. The concordance for identifying coding MMR gene somatic mutations was high between the targeted MMR gene sequencing and WGS/WES, however, the additional information gained from WGS/WES in the five tumors tested provided clinically useful insights into the tumor aetiology and cause of MMR-deficiency. A limitation of the study is the possibility that there are genes outside of those included in our 3 Tiers that influence MMR gene and/or protein expression. For example, overexpression of miR-155 has been shown to significantly down-regulate the MMR proteins, inducing a mutator phenotype and MSI^46^. Furthermore, we were unable to explore potential somatic mosaicism, which although rare, has been previously reported in individuals with Lynch syndrome^11, 47, 48^.

## CONCLUSION

In this study, we assessed the utility of WGS as a broader diagnostic tool for the detection of germline pathogenic variants in a cohort of 14 SLS cases, identifying three carriers of MMR pathogenic variants including a 9.5 Mb inversion in multiple family members. Our study further highlights the diagnostic benefit of tumor sequencing of the MMR genes in SLS cases. We identified double somatic MMR gene mutations, and therefore a likely sporadic aetiology, as the cause of tumor MMR-deficiency for >70% of the SLS cases in this study (excluding the three identified LS carriers). Additionally, WGS or WES of the SLS tumors provided additional interrogation for LOH and enabled determination of tumor MMR-deficiency related features of MSI, tumor mutational burden and somatic mutational signatures. The combined analysis of germline and tumor WGS in a SLS mother-daughter pair provided evidence against Lynch syndrome as the cause of tumor MMR-deficiency in these two relatives. As costs of high-throughput DNA sequencing continues to fall, the application of a tumor sequencing approach has the potential to replace the current LS screening methodology based on tumor immunohistochemistry, PCR-based MSI analysis and germline multigene panel sequencing and has been supported recently^49^. Testing for double somatic MMR mutations is currently not part of routine clinical practice, and is not part of the recommendations of the American College of Medical Genetics and Genomics guidelines for genetic testing for Lynch syndrome.

## Data Availability

Data is available upon request

## Grant support

Funding by a National Health and Medical Research Council of Australia (NHMRC) project grant 1125269 (PI-Daniel Buchanan), supported the design, analysis and interpretation of data. DDB is supported by a NHMRC R.D. Wright Career Development Fellowship and funding from the University of Melbourne Research at Melbourne Accelerator Program (R@MAP). BJP is supported by a Victorian Health and Medical Research Fellowship from the Department of Health and Human Services in the State of Victoria. PG is supported by an Australian Government Research Training Program Scholarship. ABS was supported by an NHMRC Senior Research Fellowship (ID1061779).

“Research reported in this publication was supported by the National Cancer Institute of the National Institutes of Health under Award Number UM1CA167551 and through cooperative agreements with the following CCFR centers: Australasian Colorectal Cancer Family Registry (NCI/NIH U01 CA074778 and U01/U24 CA097735) and by the Victorian Cancer Registry, Australia. This research was performed under CCFR approved project C-AU-1014-01.

## Financial disclosure

Dr Buchanan served as a consultant on the Tumor Agnostic (dMMR) Advisory Board of Merck Sharp and Dohme in 2017 and 2018 for Pembrolizumab.

## Acknowledgments

We thank members of the Colorectal Oncogenomics Group for their support of this manuscript. We thank the participants and staff from the Colon-CFR in particular, Maggie Angelakos, Samantha Fox and Allyson Templeton for their support of this manuscript. Computation and bioinformatics was provided by Melbourne Bioinformatics on its Peak Computing Facility.

“The content of this manuscript does not necessarily reflect the views or policies of the National Cancer Institute or any of the collaborating centers in the Colon Cancer Family Registry (Colon-CFR), nor does mention of trade names, commercial products, or organizations imply endorsement by the US Government or the Colon-CFR.”

## Author contributions

DDB, MC and CR conceived the original study concept and design and designed the analysis. The sample curation and laboratory testing was performed by MC, JJ, RW, JC, SP. BJP, KM and PG implemented the bioinformatics analysis pipeline software. BP, KM, MC and DDB prepared the manuscript. RH, SJ, AKW, ABS, FAM, IMW, JLH, MAJ contributed to the acquisition of study data. All authors provided critical revisions to the manuscript for important intellectual content have read and approved of the final manuscript.

### >Abbreviations

WGS: (whole genome sequencing)
WES: (whole exome sequencing)
MMR: (mismatch repair)
IHC: (immunohistochemical)
MSI: (microsatellite instability)
VCF: (variant call format)
LS: (Lynch syndrome)
SLS: (suspected Lynch Syndrome)
CRC: (colorectal cancer)
EC: (endometrial cancer)
MLPA: (multiplex ligation-dependent PCR amplification)
SV: (structural variant)
SNV: (single nucleotide polymorphism)
INDEL: (insertion or deletion)
PCR: (polymerase chain reaction)
HTS: (High Throughput Sequencing)
VAF: (variant allele fraction)
ACCFR: (Australasian Colorectal Cancer Family Registry).

## REFERENCES

1. Ligtenberg MJ, Kuiper RP, Chan TL, et al. Heritable somatic methylation and inactivation of MSH2 in families with Lynch syndrome due to deletion of the 3’ exons of TACSTD1. Nat Genet 2009;41:112–7.

2. Win AK, Young JP, Lindor NM, et al. Colorectal and other cancer risks for carriers and noncarriers from families with a DNA mismatch repair gene mutation: a prospective cohort study. J Clin Oncol 2012;30:958–64.

3. Thibodeau SN, Bren G, Schaid D. Microsatellite instability in cancer of the proximal colon. Scienc. 1993;260:816–9.

4. Buchanan DD, Rosty C, Clendenning M, et al. Clinical problems of colorectal cancer and endometrial cancer cases with unknown cause of tumor mismatch repair deficiency (suspected Lynch syndrome). Appl Clin Genet 2014;7:183–93.

5. Klarskov L, Ladelund S, Holck S, et al. Interobserver variability in the evaluation of mismatch repair protein immunostaining. Hum Pathol 2010;41:1387–96.

6. Markow M, Chen W, Frankel WL. Immunohistochemical Pitfalls: Common Mistakes in the Evaluation of Lynch Syndrome. Surg Pathol Clin 2017;10:977–1007.

7. Boland CR. The mystery of mismatch repair deficiency: lynch or lynch-like? Gastroenterology 2013;144:868–70.

8. Clendenning M, Buchanan DD, Walsh MD, et al. Mutation deep within an intron of MSH2 causes Lynch syndrome. Fam Cancer 2011;10:297–301.

9. Morak M, Heidenreich B, Keller G, et al. Biallelic MUTYH mutations can mimic Lynch syndrome. Eur J Hum Genet 2014.

10. Elsayed FA, Kets CM, Ruano D, et al. Germline variants in POLE are associated with early onset mismatch repair deficient colorectal cancer. Eur J Hum Genet 2015;23:1080–4.

11. Sourrouille I, Coulet F, Lefevre JH, et al. Somatic mosaicism and double somatic hits can lead to MSI colorectal tumors. Fam Cancer 2013;12:27–33.

12. Mensenkamp AR, Vogelaar IP, van Zelst-Stams WA, et al. Somatic mutations in MLH1 and MSH2 are a frequent cause of mismatch-repair deficiency in Lynch syndrome-like tumors. Gastroenterolog. 2014;146:643–646 e8.

13. Haraldsdottir S, Hampel H, Tomsic J, et al. Colon and endometrial cancers with mismatch repair deficiency can arise from somatic, rather than germline, mutations. Gastroenterolog. 2014;147:1308–1316 e1.

14. Pearlman R, Haraldsdottir S, de la Chapelle A, et al. Clinical characteristics of patients with colorectal cancer with double somatic mismatch repair mutations compared with Lynch syndrome. J Med Genet 2019;56:462–470.

15. Newcomb PA, Baron J, Cotterchio M, et al. Colon Cancer Family Registry: an international resource for studies of the genetic epidemiology of colon cancer. Cancer Epidemiol Biomarkers Prev 2007;16:2331–43.

16. Jenkins MA, Win AK, Templeton AS, et al. Cohort Profile: The Colon Cancer Family Registry Cohort (CCFRC). Int J Epidemiol 2018.

17. Buchanan DD, Tan YY, Walsh MD, et al. Tumor mismatch repair immunohistochemistry and DNA MLH1 methylation testing of patients with endometrial cancer diagnosed at age younger than 60 years optimizes triage for population-level germline mismatch repair gene mutation testing. J Clin Oncol 2014;32:90–100.

18. Vasen HF, Watson P, Mecklin JP, et al. New clinical criteria for hereditary nonpolyposis colorectal cancer (HNPCC, Lynch syndrome) proposed by the International Collaborative group on HNPCC. Gastroenterolog. 1999;116:1453–6.

19. Umar A, Boland CR, Terdiman JP, et al. Revised Bethesda Guidelines for hereditary nonpolyposis colorectal cancer (Lynch syndrome) and microsatellite instability. J Natl Cancer Inst 2004;96:261–8.

20. Buchanan DD, Clendenning M, Rosty C, et al. Tumor testing to identify lynch syndrome in two Australian colorectal cancer cohorts. J Gastroenterol Hepatol 2017;32:427–438.

21. Lindor NM, Burgart LJ, Leontovich O, et al. Immunohistochemistry versus microsatellite instability testing in phenotyping colorectal tumors. J Clin Oncol 2002;20:1043–8.

22. Weisenberger DJ, Siegmund KD, Campan M, et al. CpG island methylator phenotype underlies sporadic microsatellite instability and is tightly associated with BRAF mutation in colorectal cancer. Nat Genet 2006;38:787–93.

23. Buchanan DD, Sweet K, Drini M, et al. Risk factors for colorectal cancer in patients with multiple serrated polyps: a cross-sectional case series from genetics clinics. PLoS ONE 2010;5:e11636.

24. Campbell BB, Light N, Fabrizio D, et al. Comprehensive Analysis of Hypermutation in Human Cancer. Cel. 2017;171:1042–1056 e10.

25. Fishilevich S, Nudel R, Rappaport N, et al. GeneHancer: genome-wide integration of enhancers and target genes in GeneCards. Database (Oxford) 2017;2017.

26. Seifert BA, McGlaughon JL, Jackson SA, et al. Determining the clinical validity of hereditary colorectal cancer and polyposis susceptibility genes using the Clinical Genome Resource Clinical Validity Framework. Genet Med 2019;21:1507–1516.

27. Adam R, Spier I, Zhao B, et al. Exome Sequencing Identifies Biallelic MSH3 Germline Mutations as a Recessive Subtype of Colorectal Adenomatous Polyposis. Am J Hum Genet 2016;99:337–51.

28. Weren RD, Ligtenberg MJ, Kets CM, et al. A germline homozygous mutation in the base-excision repair gene NTHL1 causes adenomatous polyposis and colorectal cancer. Nat Genet 2015;47:668–71.

29. Grolleman JE, de Voer RM, Elsayed FA, et al. Mutational Signature Analysis Reveals NTHL1 Deficiency to Cause a Multi-tumor Phenotype. Cancer Cell 2019;35:256–266 e5.

30. Knijnenburg TA, Wang L, Zimmermann MT, et al. Genomic and Molecular Landscape of DNA Damage Repair Deficiency across The Cancer Genome Atlas. Cell Rep 2018;23:239–254 e6.

31. Wood RD, Mitchell M, Lindahl T. Human DNA repair genes, 2005. Mutat Res 2005;577:275–83.

32. Rhees J, Arnold M, Boland CR. Inversion of exons 1-7 of the MSH2 gene is a frequent cause of unexplained Lynch syndrome in one local population. Fam Cancer 2013.

33. Vargas-Parra GM, Gonzalez-Acosta M, Thompson BA, et al. Elucidating the molecular basis of MSH2-deficient tumors by combined germline and somatic analysis. Int J Cancer 2017;141:1365–1380.

34. Salvador MU, Truelson MRF, Mason C, et al. Comprehensive Paired Tumor/Germline Testing for Lynch Syndrome: Bringing Resolution to the Diagnostic Process. J Clin Oncol 2019;37:647–657.

35. Jansen AM, Geilenkirchen MA, van Wezel T, et al. Whole Gene Capture Analysis of 15 CRC Susceptibility Genes in Suspected Lynch Syndrome Patients. PLoS One 2016;11:e0157381.

36. Geurts-Giele WR, Leenen CH, Dubbink HJ, et al. Somatic aberrations of mismatch repair genes as a cause of microsatellite-unstable cancers. J Pathol 2014;234:548–59.

37. Wagner A, van der Klift H, Franken P, et al. A 10-Mb paracentric inversion of chromosome arm 2p inactivates MSH2 and is responsible for hereditary nonpolyposis colorectal cancer in a North-American kindred. Genes Chromosomes Cancer 2002;35:49–57.

38. Mork ME, Rodriguez A, Taggart MW, et al. Identification of MSH2 inversion of exons 1-7 in clinical evaluation of families with suspected Lynch syndrome. Fam Cancer 2017;16:357–361.

39. Shia J, Zhang L, Shike M, et al. Secondary mutation in a coding mononucleotide tract in MSH6 causes loss of immunoexpression of MSH6 in colorectal carcinomas with MLH1/PMS2 deficiency. Mod Pathol 2013;26:131–8.

40. Alexandrov LB, Nik-Zainal S, Wedge DC, et al. Signatures of mutational processes in human cancer. Natur. 2013;500:415–21.

41. Haradhvala NJ, Kim J, Maruvka YE, et al. Distinct mutational signatures characterize concurrent loss of polymerase proofreading and mismatch repair. Nat Commun 2018;9:1746.

42. Castillejo A, Vargas G, Castillejo MI, et al. Prevalence of germline MUTYH mutations among Lynch-like syndrome patients. Eur J Cancer 2014;50:2241–50.

43. Jansen AM, van Wezel T, van den Akker BE, et al. Combined mismatch repair and POLE/POLD1 defects explain unresolved suspected Lynch syndrome cancers. Eur J Hum Genet 2016;24:1089–92.

44. Xavier A, Olsen MF, Lavik LA, et al. Comprehensive mismatch repair gene panel identifies variants in patients with Lynch-like syndrome. Mol Genet Genomic Med 2019:e850.

45. Keijzers G, Liu D, Rasmussen LJ. Exonuclease 1 and its versatile roles in DNA repair. Crit Rev Biochem Mol Biol 2016;51:440–451.

46. Valeri N, Gasparini P, Fabbri M, et al. Modulation of mismatch repair and genomic stability by miR-155. Proc Natl Acad Sci U S A 2010;107:6982–7.

47. Pastrello C, Fornasarig M, Pin E, et al. Somatic mosaicism in a patient with Lynch syndrome. Am J Med Genet A 2009;149A:212–5.

48. Geurts-Giele WR, Rosenberg EH, Rens AV, et al. Somatic mosaicism by a de novo MLH1 mutation as a cause of Lynch syndrome. Mol Genet Genomic Med 2019;7:e00699.

49. Hampel H, Pearlman R, Beightol M, et al. Assessment of Tumor Sequencing as a Replacement for Lynch Syndrome Screening and Current Molecular Tests for Patients With Colorectal Cancer. JAMA Oncol 2018;4:806–813.

